# Teacher- and Parent-Reported Trajectories of Challenging Behavior Among Individuals with Autism

**DOI:** 10.1101/2022.09.09.22279781

**Authors:** Nicole E. Rosen, Hillary K. Schiltz, Catherine Lord

## Abstract

People with autism spectrum disorder (ASD) frequently exhibit challenging behaviors throughout the lifespan, which can have pervasive effects on quality of life. Challenging behaviors have been shown to change over time as a function of various individual-level factors (e.g., cognitive ability), yet research is primarily limited to parent-reported measures. To expand upon this work, the present study aimed to examine trajectories of teacher- and parent-reported challenging behaviors (i.e., hyperactivity, irritability, social withdrawal) and to test whether predictors including ASD features, verbal intelligence quotient, and consistency in reporter impact these trajectories among individuals with ASD or non-spectrum delays from ages 9 to 18. Multilevel models revealed that, according to both teacher and parent report, participants showed the greatest improvement in hyperactivity, less but still notable improvement in irritability, and stable levels of social withdrawal over time. Higher cognitive ability and fewer ASD features emerged as important individual differences related to fewer challenging behaviors. The multi-informant perspective and longitudinal design provide novel insight into the manifestations of these challenging behaviors across different contexts and across time. Findings highlight the importance of addressing challenging behaviors as these behaviors tend to persist throughout development in both home and school contexts, especially for children with particular diagnostic and cognitive profiles.

**Lay Summary:** According to both teacher and parent report, youth with autism showed the greatest improvement in hyperactivity, less but still notable improvement in irritability, and stable levels of social withdrawal from school-age to adolescence. Fewer autism features and greater cognitive ability were related to fewer challenging behaviors. This study’s use of multiple reporters (e.g., teachers and parents) across time provided insight into the persistence of challenging behaviors in the home and school settings and across development.

---

Individuals with autism spectrum disorder (ASD) can have challenging or maladaptive behaviors throughout the lifespan (Lecavalier, 2006; Shattuck et al., 2007; Simonoff et al., 2008). Challenging behaviors interfere with everyday functioning and include behaviors such as hyperactivity, irritability, social withdrawal, self-injury, and aggression (Shattuck et al., 2007). These behaviors are more common among children with ASD than among children with other conditions or children without ASD (Blacher & McIntyre, 2006; Gadow et al., 2005; Gillot et al., 2001; Tonge & Einfield, 2003), with studies identifying a prevalence rate of at least 50% in ASD (Baghdadli et al., 2003; Holden & Gitlesen, 2006; Kozlowski & Matson, 2012; Matson et al., 2009; Simonoff et al., 2008). Challenging behaviors have been shown to affect academic achievement, social competence, and adult psychiatric functioning both among individuals with and without ASD (de Bildt et al., 2005; Kim et al., 2000) and strongly impact caregiver and teacher stress (Lecavalier et al., 2006). However, despite the high prevalence and functional impairment associated with challenging behaviors in ASD, our understanding of how these behaviors change across time and across contexts consists primarily of parent-report measures administered cross-sectionally. In the present study, we take advantage of longitudinal data from multiple informants (e.g., parents and teachers) to begin to address these questions.

Challenging behaviors in ASD may wax and wane across development due to a host of external (e.g., social contexts) and internal (e.g., biological and developmental) factors. For example, the increased independence, more nuanced social interactions, and higher social expectations and demands that accompany adolescence, coupled with the onset of puberty, may exacerbate maladaptive behaviors including social withdrawal, irritability, aggression, and destructiveness (Anderson et al., 2011; Kobayashi et al., 1992). At the same time, self-regulatory abilities become increasingly internalizing (Sameroff, 2010) as the frontal lobe and associated executive function abilities develop (Blakemore & Choudhury, 2006), which sets the stage for improved self-control over challenging behaviors as a child matures. Therefore, longitudinal research on challenging behaviors is critical to shed light on how these behaviors unfold across childhood and adolescence in ASD.

Although there are few longitudinal studies tracking challenging behavior trajectories in ASD over time, current findings based on parent report suggest improvement in challenging behaviors with age, particularly externalizing behaviors (Shattuck et al., 2007; Stringer et al., 2020; Woodman et al., 2015). This work includes a parent-reported study of participants who overlap with the current sample (Anderson et al., 2011). However, improvement is not universal; for example, cognitive ability is consistently identified as a significant predictor of challenging behavior trajectories, such that individuals with ASD with lower cognitive ability not only present with higher levels of challenging behaviors, but also experience slower improvement in these behaviors over time (Anderson et al., 2011; Shattuck et al., 2007; Woodman et al., 2015). Greater autism features have also been found to be associated with higher levels of challenging behaviors (Anderson et al., 2011; Matson et al., 2009), though the impact of ASD features on patterns of challenging behavior in ASD over time is not well-studied and findings have been mixed. For example, some studies have found no significant relationship between the severity of ASD and challenging behavior trajectories (Stringer et al., 2020), while others have found an association between greater severity of ASD and greater improvement in specific types of challenging behavior, namely hyperactivity and irritability, with less improvement in social withdrawal (Anderson et al., 2011). No research to the authors’ knowledge, however, has analyzed challenging behavior trajectories in ASD from perspectives outside of parent report (Anderson et al., 2011; Gray et al., 2012; Stringer et al., 2020), and therefore it is unknown whether these patterns replicate across various reporters and contexts.

Teachers, in particular, can provide unique insight into how challenging behaviors manifest in children with ASD across varied situations and contexts outside of those observed by parents, including in the classroom and amongst peers. The complexity of ASD, variability in behaviors observed across contexts, and impact of the environment on reporting (De Los Reyes & Kazdin, 2005; Dickson et al., 2018; Kraemer et al., 2003) further support the need for a multi-informant approach to research and assessment in ASD. Additionally, informants often differ in how they interact with the individual with ASD as well as how the presence of the informant influences behavior (De Los Reyes, 2011). Overall, evidence points to low-to-moderate agreement between caregiver and teacher reports of challenging behaviors in ASD (McDonald et al., 2016; Stadnick et al., 2017; Stratis & Lecavalier, 2015). While parent-report trajectories of challenging behaviors in ASD have been documented (Anderson et al., 2011; Matson et al., 2009; Shattuck et al., 2007; Stringer et al., 2020), the important perspective of teachers has yet to be captured over time.

Although we have some understanding of the challenging behavior trajectories and individual-level factors that influence them from the parent perspective, it is important to concurrently assess patterns from other perspectives, especially teachers. In light of this need, the present study aimed to 1) examine trajectories of teacher- and parent-reported challenging behaviors (i.e., hyperactivity, irritability, social withdrawal), and 2) test whether individual- and teacher-level predictors (i.e., autism features, verbal intelligence quotient (VIQ), consistency in reporter, teacher training, student-teacher ratio) impact these trajectories among individuals with ASD or non-spectrum delays from ages 9 to 18.

## Methods

### Participants

Participants were recruited from three sources: (1) 192 children under age 3 years referred for possible ASD to two tertiary autism programs (North Carolina and Illinois); (2) 21 children under age 3 years with non-ASD developmental delays identified through the referral sources of the first group (North Carolina and Illinois); and (3) 40 children with ASD or neurodevelopmental delays also diagnosed at early ages who joined the study at approximately age 9 and then were followed at the same ages as the first two groups (Michigan) (see Anderson et al., 2014 for additional recruitment details). By age 9 (mean (*M*) years=9.98, standard deviation (*SD*)=0.89), the full sample had joined the study and most had completed one or more in-person assessments. Additional in-person assessments occurred at approximately ages 19 (*M*=19.04, *SD*=1.2) and 26 (*M*=25.97, *SD*=1.4). Biannual packets of questionnaires were completed throughout the entirety of the study.

Of the original 253 participants, 165 were selected for this study based on availability of teacher- and parent-reported data between ages 9 to 18. A majority had more than one time point of teacher-reported data (77.6% completed two or more, 70.9% completed three or more, 63.6% completed four or more, 57% completed five or more) and parent-reported data (86.1% completed two or more, 83% completed three or more, 80% completed four or more, 77.6% completed five or more), with over five teacher-reported time points and over ten parent-reported time points on average per participant. Attrition in our sample was higher among participants with lower levels of autism features based on Autism Diagnostic Observation Schedule (ADOS; Lord et al., 2000, 2012) calibrated severity scores (*p*=.035) and more advanced caregiver education (*p*=.006), but not associated with gender, recruitment site, diagnosis, race, or VIQ. Among the 165 participants in the current sample, 24.8% identified as Black and the remainder identified as White (see Table 1). The sample was predominantly male (79.4%) with 52.1% from North Carolina, 30.9% from Illinois, and 17% from Michigan. Approximately half of the sample reported a caregiver education level of at least a 4-year college degree (47.9%). Additionally, 21.2% of the sample never received a formal diagnosis of ASD throughout the course of the longitudinal study. These participants are included in this study because they show similar patterns in presentation and outcome across development to the participants with ASD (see Lord et al., 2020; McCauley et al., 2020); to account for these diagnostic differences, the ASD history of participants (ever/never ASD) was tested as a covariate.

**Table 1.** Sample demographic and diagnostic characteristics.

|  | (*N* = 165) |
| --- | --- |
| **Male, *n* (%)** | 131 (79.4) |
| **White, *n* (%)** | 124 (75.2) |
| **Caregiver education** |  |
| **College degree or above, *n* (%)** | 79 (47.9) |
| **Site, *n* (%)** |  |
| **North Carolina** | 86 (52.1) |
| **Illinois** | 51 (30.9) |
| **Michigan** | 28 (17) |
| **ASD dx, *n* (%)** | 130 (78.8) |
| **ADOS-CSS, *M* (*SD*)** | 6.18 (2.84) |
| **VIQ, *M* (*SD*)** | 56.42 (37.5) |

ASD dx: autism spectrum disorder diagnosis (diagnostic history); ADOS-CSS: Autism Diagnostic Observation Schedule-Calibrated Severity Score; VIQ: verbal intelligence quotient; *M*: mean; *SD*: standard deviation

### Procedures

This research was approved by institutional review boards at various institutions. Informed consent was obtained from all caregivers and individuals themselves whenever possible. Various diagnostic instruments, parent interviews, cognitive testing, questionnaires, and standard demographic forms were administered in-person (see Anderson et al., 2014). Participants, their parents, and their teachers also completed questionnaires via mail multiple times throughout the study (16 times spaced on average 6 months apart between ages 9-18). Teacher packets, which included teacher consents, questionnaires, and pre-addressed return envelopes, were mailed to parents with the instruction to distribute to the identified teacher/s. Teachers were mailed a small gift as compensation. If the teacher forms were not returned, parents consented for teachers to be contacted directly by the study team. Clinicians conducting the in-person assessments, generally a post-doctoral fellow or licensed clinician and a research assistant, were research reliable in the relevant measures and were blind to the participants’ previous assessment results. Diagnoses of ASD or other disorders were made by the research team and presented to a panel of experienced clinicians to come to a consensus on diagnoses of ASD and other conditions. All assessments were provided free of charge and included feedback on testing results.

### Measures

#### Autism Features

Participant ADOS-Calibrated Severity Score (ADOS-CSS) at age 9 (if unavailable, then from later years) was used to describe level of autism features. At each in-person assessment at approximately ages 9, 19, and 26 years, participants were administered the ADOS (Lord et al., 2000, 2012). The ADOS-CSS (Gotham et al., 2009) can be used to compare ASD features across individuals of different developmental levels. ADOS-CSS scores range from 1 to 10, with higher scores indicating higher levels of autism spectrum–related features observed during the ADOS.

#### Cognitive Abilities

Cognitive assessments were administered at each face-to-face assessment. The instrument used to obtain VIQ scores at age 9 (if unavailable, from later years) was chosen from a standard hierarchy including the Wechsler Abbreviated Scale of Intelligence (WASI; Wechsler, 1999), Wechsler Intelligence Scale for Children (WISC-III; Wechsler, 1991), and Differential Abilities Scale (DAS; Elliott, 1990, 2007), depending on the child’s language level and previous cognitive testing battery. Ratio VIQs were calculated from age equivalents when raw scores were outside deviation score ranges. VIQ, as opposed to nonverbal intelligence quotient, was used for sake of consistency with existing literature.

#### Challenging Behavior

The Aberrant Behavior Checklist (ABC; Aman et al., 1985) is a 58-item checklist designed to assess challenging behaviors in individuals with developmental disabilities across five subscales: Hyperactivity, Irritability, Social Withdrawal/Lethargy, Stereotypic Behavior, and Inappropriate Speech. Items are scored on a four-point Likert scale ranging from (0) “not at all a problem” to (3) “the problem is severe in degree”. For this study, the Stereotypic Behavior and Inappropriate Speech subscales were excluded from analyses due to the significant overlap of these items with core ASD features (Anderson et al., 2011; Fok & Bal, 2019). Furthermore, hyperactivity, irritability, and social withdrawal/lethargy emerge as particularly important outcomes in ASD given their high prevalence (Simonoff et al., 2008) and significant impact on quality of life (Anderson et al., 2011; Gerber et al., 2008). Teachers and parents completed the ABC yearly to capture participants’ challenging behaviors from approximately ages 9 to 18. Teachers also reported whether they were a special or general education teacher and the number of students and staff in their classroom to calculate student-adult ratios in the classroom. The ABC has been found to have robust psychometric properties including moderate-to-high reliability (Aman et al., 1987), good validity (Aman et al., 1985; Rojahn & Helsel, 1991), and strong psychometric performance among individuals with ASD (Brinkley et al., 2006; Kaat et al., 2014; Norris et al., 2019).

### Statistical Analyses

Changes in teacher- and parent-reported ABC-Hyperactivity, -Irritability, and -Social Withdrawal from ages 9 to 18 were each estimated using multilevel models and maximum likelihood estimation with robust standard errors (MLR) in M*plus* v. 8.1. Our multilevel structures included age/time (level 1), individual (level 2), and site (level 3). First, null models were used to test whether random effects describing between-participant and between-recruitment site were appropriate (Finch & Bolin, 2017; Luke, 2020). Second, unconditional growth models tested the rate of change (i.e., slope) of ABC-Hyperactivity, -Irritability, and - Social Withdrawal as a function of participant age as a fixed effect. The intercept of the model was set to age 9 for interpretability. Third, we tested if random effects of age (i.e., differences in slope across people) improved model fit using a chi-square difference test based on loglikelihood values and scaling correction factors. Fourth, demographic and individual descriptive covariates including gender, race, caregiver education, ADOS-CSS, and VIQ were each tested individually as predictors of the intercept or slope (i.e., cross-level interactions with age) in the baseline models. The pattern of findings for ADOS-CSS and history of ASD was highly similar; therefore, only ADOS-CSS was retained in models to capture a more fine-grained rating of autism features. Additionally, for teacher-reported trajectories, teacher-level covariates including consistency in reporter (ratio of participant ABC entries per unique teacher; *M*=1.87, *SD*=1.18, Range=1-10), student-adult ratio (ratio of the number of students per adults in the classroom; *M*=5.69, *SD*=4.19, Range=1-40), and teacher type (special or general education teacher; 121 (73.3%) special education teachers) were also tested in the baseline models. Lastly, teacher- and parent-reported models were run that included multiple predictors based on the previous analyses and tested relevant interactions between predictors (e.g., ADOS-CSS and VIQ) on intercept and slope of hyperactivity, irritability, and social withdrawal from ages 9 to 18. Predictor variables were mean-centered for ease of interpretability and plotted in graphs one standard deviation above or below the mean.

## Results

### Primary Baseline Model Analyses

The null model without predictors revealed that significant variation in ABC-Hyperactivity, ABC-Irritability, and ABC-Social Withdrawal could be attributed to between-participant differences for teacher-reported (intraclass correlation coefficient (ICC)=0.523, 0.531, 0.444, respectively) and parent-reported data (ICC=0.75, 0.75, 0.804, respectively), but not to between-recruitment site differences for teacher-reported (ICC=0.000, 0.001, 0.002, respectively) or parent-reported data (ICC=0.000, 0.000, 0.000, respectively). ABC teacher and parent report in the current sample show moderate interrater agreement when tested across multiple time points from ages 9 to 18 years for hyperactivity (*r*=.380-.482) and irritability (*r*=.397-.445), and low interrater agreement for social withdrawal (*r*=.097-.252).

### Hyperactivity

#### Hyperactivity Baseline Models

The baseline model showed a significant negative trajectory in ABC-Hyperactivity symptoms from late childhood into early adulthood as a function of age, indicating improvement over time according to both teacher report (*B*=-0.54, *p*<.001; see Figure 1a) and parent report (*B*=-0.66, *p*<.001; see Figure 1b). The random effect of age significantly improved model fit (*p*<.001), indicating significant variability in trajectory of hyperactivity across people based on teacher and parent report.

**Figure 1.**
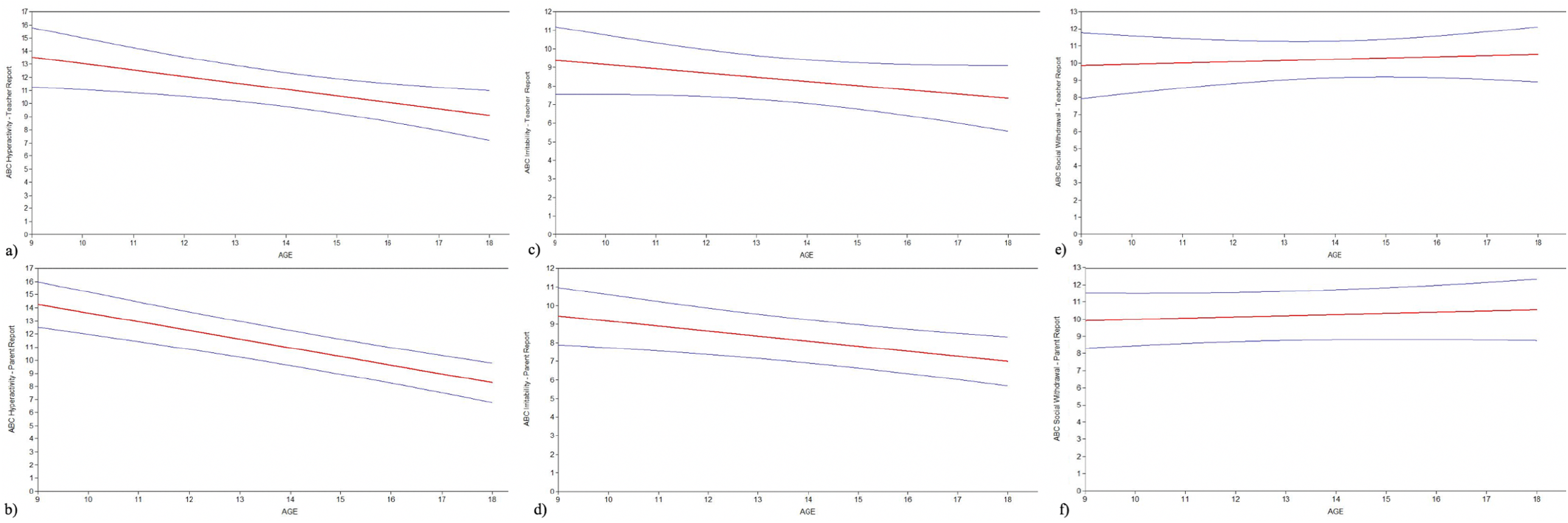
Teacher- and Parent-Reported Baseline Trajectories.

#### Predictors of Teacher-Reported Hyperactivity

When tested individually in the teacher-report models, consistency in reporter, ADOS-CSS, and VIQ all emerged as significant predictors of ABC-Hyperactivity at age 9 (intercept) or change in ABC-Hyperactivity scores from ages 9 to 18 (slope); gender, race, caregiver education, student-adult ratio, and teacher type were not significant predictors of model intercept or slope. Consistency in reporter (i.e., whether the same teacher completed the questionnaire year after year) had a nonsignificant effect on intercept (*B*=-0.78, *p*=.51) but a significant effect on slope (*B*=0.37, *p*=.03; see Figure 2a); thus, while participants presented at similar symptom levels at age 9, participants with less consistency in reporters (i.e., many different teachers reporting over time) experienced significantly greater improvement in hyperactivity across time than participants with more consistency in reporters (same reporter/s reporting multiple times). Given the clinical relevance of the overlap between ADOS-CSS and VIQ, these predictor variables were simultaneously included in the model to isolate the effect of each variable. Controlling for VIQ, ADOS-CSS had a significant effect on both intercept (*B*=0.94, *p=*.03) and slope (*B*=-0.16, *p=*.02; see Figure 2b), such that participants with higher ADOS-CSS (more ASD features) presented with significantly more hyperactivity at age 9 and experienced significantly greater improvement in hyperactivity over time than participants with lower ADOS-CSS (see Table 2). Controlling for ADOS-CSS, VIQ had a marginal effect on intercept (*B*=-0.05, *p=*.08) and a significant effect on slope (*B*=-0.01, *p=*.04; see Figure 2c), suggesting that participants with lower VIQs presented with more hyperactivity at age 9 and experienced significantly less improvement in hyperactivity over time than participants with higher VIQs. The interaction between ADOS-CSS and VIQ had nonsignificant effects on both intercept and slope in the ABC-Hyperactivity model.

**Figure 2.**
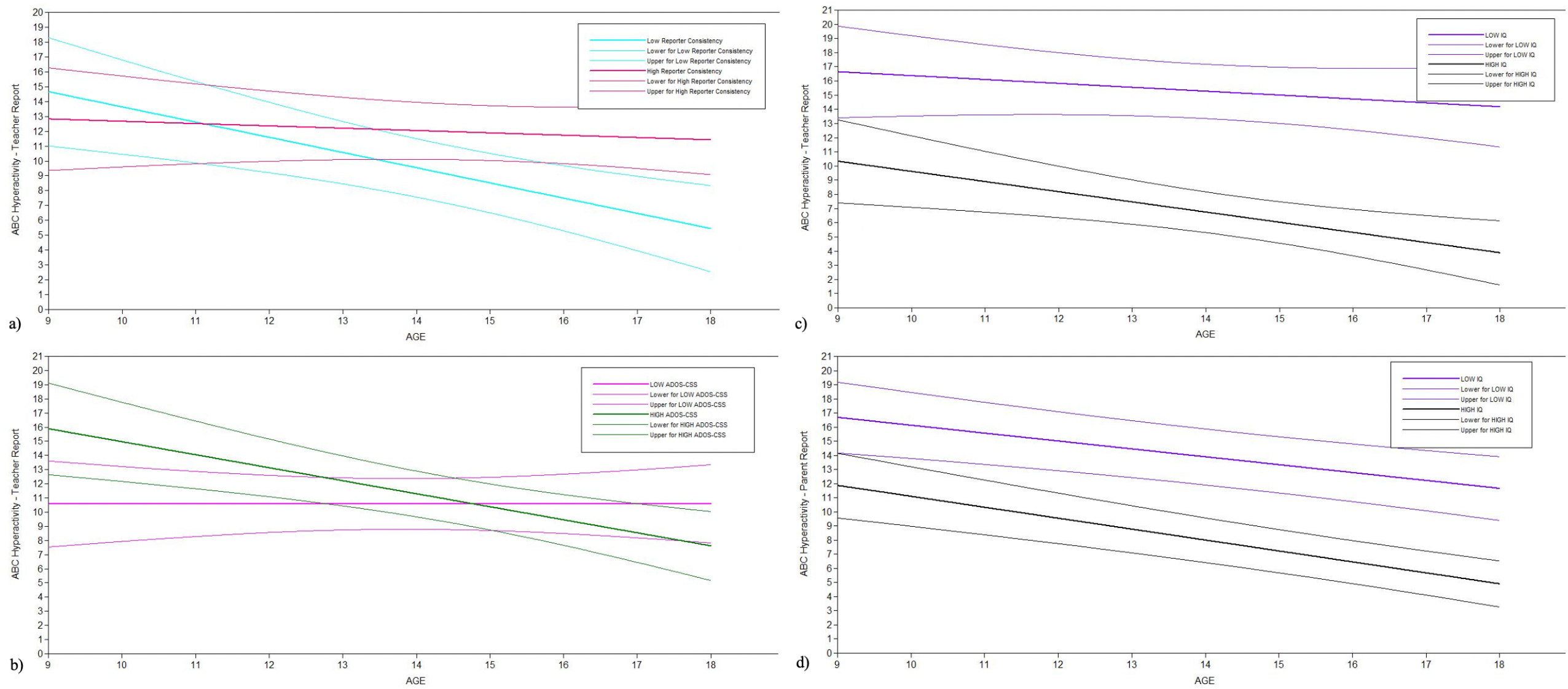
Teacher- and Parent-Reported Predictors of ABC-Hyperactivity Trajectories.

**Table 2.**
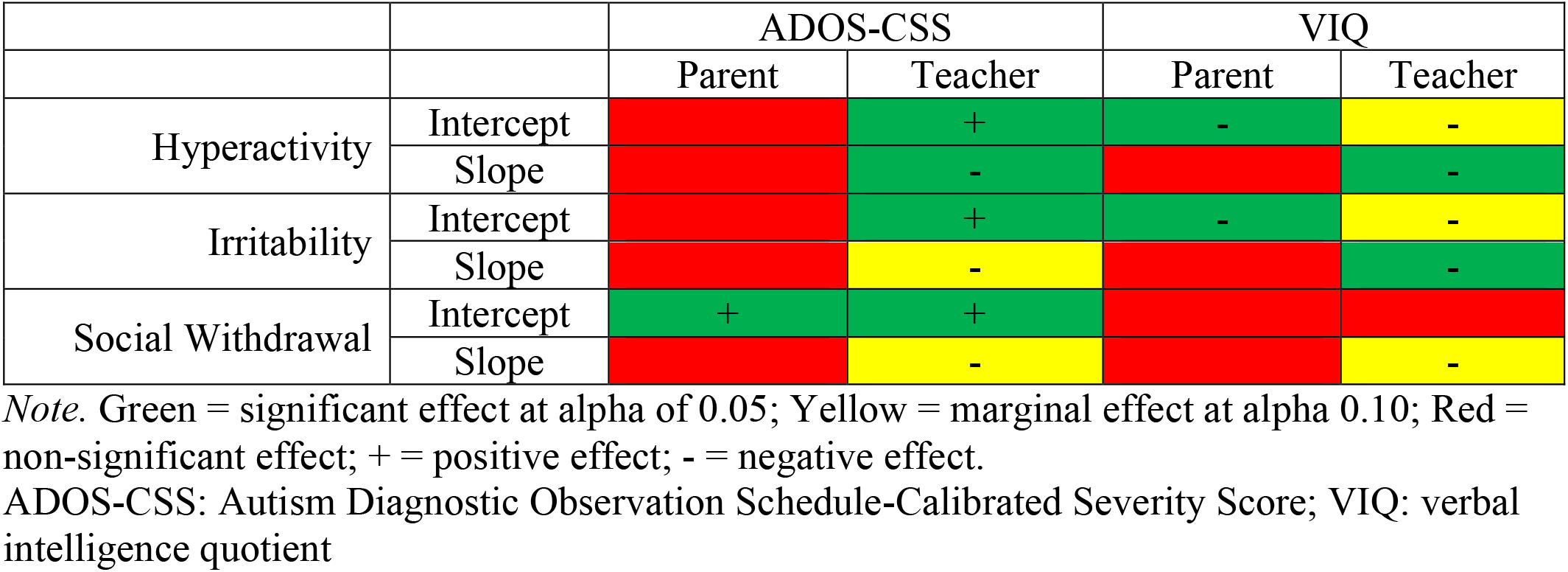
Predictor effects on ABC trajectories.

#### Predictors of Parent-Reported Hyperactivity

When predictors were tested individually in the parent-report ABC-Hyperactivity model, VIQ had a significant inverse effect on intercept (*B*=-0.07, *p*=.01), but a nonsignificant effect on slope (*B*=-0.003, *p*=.27; see Figure 2d). Consistent with teacher report, while participants with lower VIQs presented with significantly more hyperactivity at age 9 than participants with higher VIQs, but in contrast to teacher report, participants with higher and lower VIQs did not differ in their rate of improvement in hyperactivity across time. Gender, race, and caregiver education were not significant predictors of parent-reported ABC-Hyperactivity intercept or slope. Additionally, in contrast with findings for teacher report, ADOS-CSS was also not a significant predictor in the parent-reported ABC-Hyperactivity model.

### Irritability

#### Irritability Baseline Models

The baseline model showed a negative trajectory in ABC-Irritability symptoms from late childhood into early adulthood as a function of age, indicating evidence for improvement over time according to both teacher report (*B*=-0.23, *p*=.14; see Figure 1c) and parent report (*B*=-0.27, *p*=.004; see Figure 1d). A significant random effect of age was identified for both teacher and parent report (*p*<.001), indicating differences in trajectories across people.

#### Predictors of Teacher-Reported Irritability

Both participant ADOS-CSS and VIQ had significant effects on teacher-reported ABC-Irritability, though gender, race, caregiver education, consistency in reporter, student-adult ratio, and teacher type did not significantly contribute to the variation in teacher-reported ABC-Irritability at age 9 (intercept) or to change in ABC-Irritability scores from ages 9 to 18 (slope). Again, given the clinically-relevant overlap between ADOS-CSS and VIQ, these predictor variables were simultaneously included in the model to isolate the effect of each variable. Controlling for VIQ, ADOS-CSS had a significant effect on intercept (*B*=0.88, *p=*.01) and a marginal effect on slope (*B*=-0.113, *p=*.06; see Figure 3a), such that participants with higher ADOS-CSS (more ASD features) presented with significantly more irritability at age 9 and experienced slightly greater improvement in irritability over time than participants with lower ADOS-CSS. Accounting for differences in ADOS-CSS, VIQ had a marginal effect on intercept (*B*=-0.05, *p=*.06) and a significant effect on slope (*B*=-0.01, *p=*.03; see Figure 3b), suggesting that participants with lower VIQs presented with marginally more irritability at age 9 and experienced significantly less improvement in irritability over time than participants with higher VIQ. The interaction between ADOS-CSS and VIQ had nonsignificant effects on both intercept and slope in the ABC-Irritability model.

**Figure 3.**
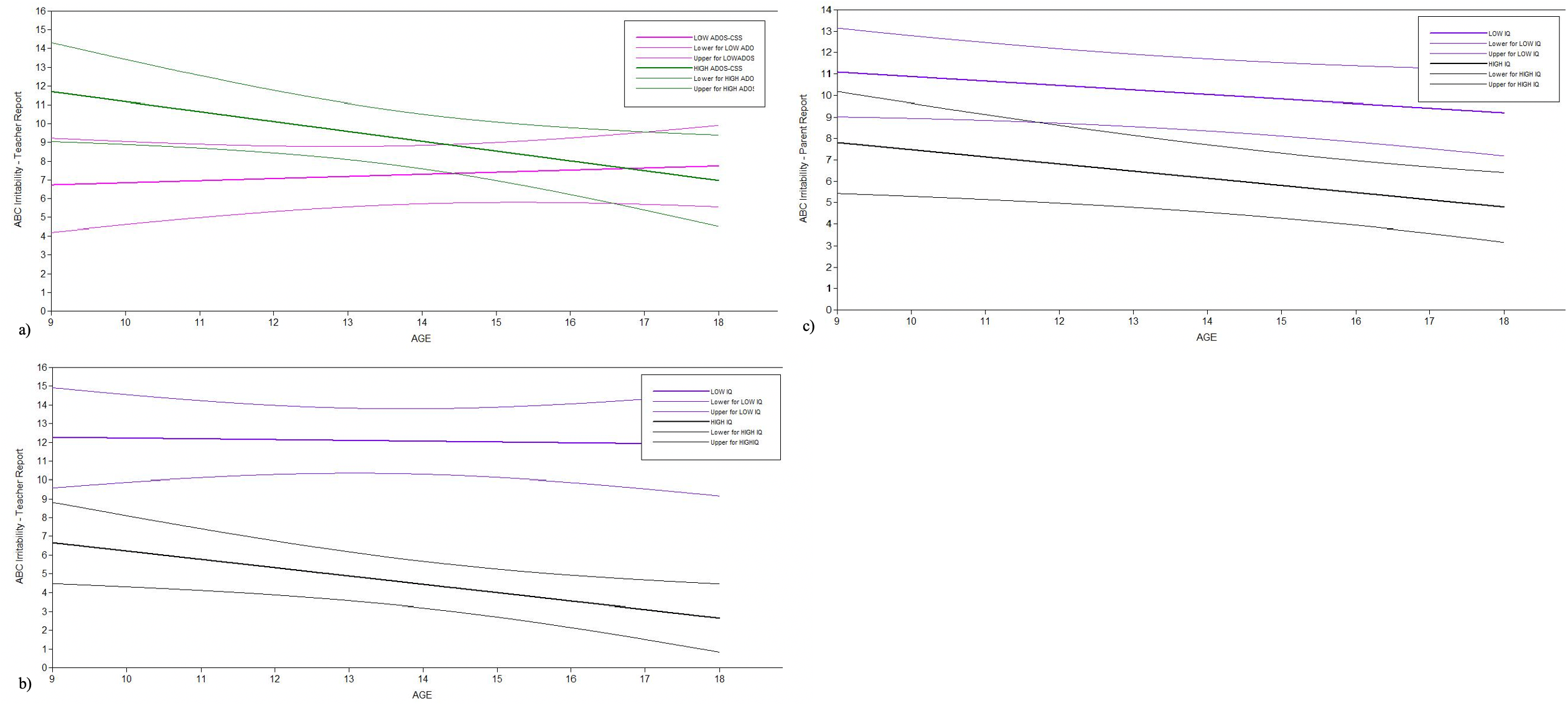
Teacher- and Parent-Reported Predictors of ABC-Irritability Trajectories.

#### Predictors of Parent-Reported Irritability

When predictors were tested individually in the parent-report models, VIQ emerged as a significant predictor in the ABC-Irritability model. Similar to teacher-reported findings, VIQ had a significant effect on intercept (*B*=-0.04, *p*=.049), but in contrast to teacher-reported findings, VIQ had a nonsignificant effect on slope (*B*=-0.002, *p*=.52; see Figure 3c); while participants with lower VIQs presented with significantly more irritability at age 9 than participants with higher VIQs, participants did not differ in their rate of improvement in irritability across time. Gender, race, and caregiver education were not significant predictors of parent-reported ABC-Irritability intercept or slope. Additionally, in contrast with findings for teacher report, ADOS-CSS was also not a significant predictor in the parent-reported ABC-Irritability model.

### Social Withdrawal

#### Social Withdrawal Baseline Models

The baseline model showed a nonsignificant slope of ABC-Social Withdrawal symptoms from late childhood into early adulthood as a function of age according to both teacher report (*B=*0.08, *p*=.64; see Figure 1e) and parent report (*B*=0.06, *p*=0.49; see Figure 1f), suggesting that social withdrawal is relatively stable across time. There was significant variability in trajectory of social withdrawal across people based on teacher and parent report (*p*<.001).

#### Predictors of Teacher-Reported Social Withdrawal

Participant ADOS-CSS and VIQ emerged as significant to marginally-significant predictors in the teacher-reported ABC-Social Withdrawal model; conversely, gender, race, caregiver education, consistency in reporter, student-adult ratio, and teacher type did not significantly contribute to variation in teacher-reported ABC-Social Withdrawal at age 9 (intercept) or to change in ABC-Social Withdrawal scores from ages 9 to 18 (slope). Per teacher report, controlling for VIQ, ADOS-CSS had a significant effect on intercept (*B*=1.40, *p*<.01) and a marginal effect on slope (*B=*-0.12, *p*=.08; see Figure 4a), such that participants with higher ADOS-CSS (more ASD features) presented with significantly more social withdrawal symptoms at age 9 and experienced marginally greater improvement over time than participants with lower ADOS-CSS. Accounting for differences in ADOS-CSS, VIQ had a nonsignificant effect on intercept (*B*=0.004, *p=*.90) and a marginal effect on slope (*B*=-0.01, *p=*.10; see Figure 4b); while participants presented with similar levels of social withdrawal symptoms at age 9 regardless of VIQ, participants with lower VIQs experienced marginally less improvement in social withdrawal symptoms over time than participants with higher VIQs.

**Figure 4.**
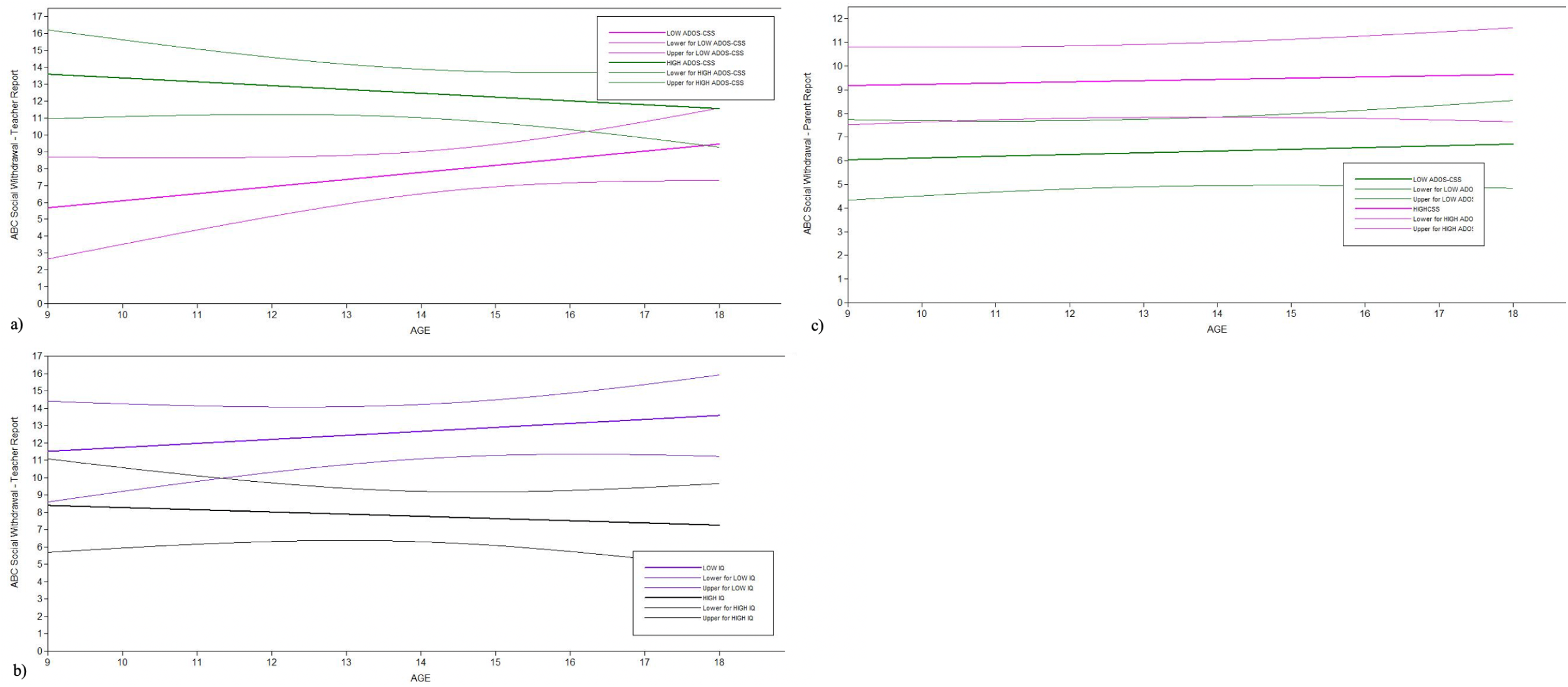
Teacher- and Parent-Reported Predictors of ABC-Social Withdrawal Trajectories.

#### Predictors of Parent-Reported Social Withdrawal

When predictors were tested individually in the parent-report models, ADOS-CSS and VIQ emerged as significant predictors in the ABC-Social Withdrawal model, though gender, race, and caregiver education failed to reach significance. Upon entering ADOS-CSS and VIQ into the model simultaneously to account for the clinical overlap, only ADOS-CSS emerged as a significant predictor of ABC-Social Withdrawal. Controlling for VIQ, ADOS-CSS had a significant effect on intercept (*B*=0.56, *p*<.01) but a nonsignificant effect on slope (*B*=-0.004, *p*=0.89; see Figure 4c); consistent with teacher report, participants with higher ADOS-CSS (more ASD features) presented with significantly more social withdrawal symptoms at age 9 than participants with lower ADOS-CSS, though, in contrast to teacher report, displayed similar rates of improvement in social withdrawal from ages 9 to 18 regardless of ADOS-CSS. Controlling for ADOS-CSS, VIQ was not a significant predictor of ABC-Social Withdrawal intercept (*B*=-0.03, *p*=0.15) or slope (*B*=-0.002, *p*=0.45). Furthermore, the interaction between ADOS-CSS and VIQ had nonsignificant effects on both intercept and slope in the ABC-Social Withdrawal model.

## Discussion

The present study extends the field’s understanding of changes in and predictors of challenging behavior trajectories among individuals with ASD and non-ASD developmental delays from both teacher and parent perspectives. Findings revealed an overall pattern of improvement from late childhood into adolescence in many, although not all, challenging behaviors. Specifically, according to both teacher and parent report, participants showed the greatest improvement in hyperactivity, less but still notable improvement in irritability, and stable levels of social withdrawal from ages 9 to 18. This is the first study, to the authors’ knowledge, to document consistency in trajectory patterns of challenging behaviors across parent and teacher report.

The improvements in hyperactivity and irritability are commensurate with the trend of improvement in behavioral functioning over time in ASD (Anderson et al., 2011, 2014; Seltzer et al., 2004; Shattuck et al., 2007; Stringer et al., 2020; Woodman et al., 2015). It may be that developmental improvements in self-regulation that come online as children age (Cibralic et al., 2019; Greenlee et al., 2021; Shattuck et al., 2007), coupled with environmental supports from parents and teachers (Beck et al., 2020; Ting & Weiss, 2017; Woodman et al., 2015), lead to better management of behaviors such as impulsivity, distractibility, tantrums, and aggression over time. The lack of improvement in social withdrawal symptoms over time broadly aligns with the existing literature. Consistent with study findings, research suggests that internalizing conditions involving social withdrawal, including depression and social anxiety, appear to peak in adolescence in ASD (Ghaziuddin et al., 2002). This peak coincides with the natural increase in the social demands of adolescence and young adulthood as youth gain more independence in social situations and receive less structure and guidance from caregivers (Hume et al., 2014; Steinberg, 2001). An analysis of a sample that overlaps with the one used for the present study found that social withdrawal at age 18 was associated with greater difficulties with friendships, increased feelings of loneliness, and more depressive symptoms (Anderson et al., 2011). Thus, the persistent nature of social withdrawal over time may be due to a bidirectional process in which individuals with ASD who are more socially withdrawn may be at greater risk for loneliness and depression, which may in turn lead to continued withdrawal.

This multi-informant study was the first, to our knowledge, to examine trajectories of challenging behaviors from both teacher and parent perspectives to provide insight into behavioral patterns across different contexts over time. Teachers and parents showed moderate interrater agreement on reports of hyperactivity and irritability symptoms, and low interrater agreement on reports of social withdrawal symptoms. Although literature on parent-teacher agreement is somewhat mixed, studies generally identify low-to-moderate correlations (Freund & Reiss, 1991; Lane et al., 2013; Stratis & Lecavalier, 2015, 2017), with better agreement on externalizing vs. internalizing constructs due to the observability of these behaviors (Stratis & Lecavalier, 2015). Perhaps most notably, teacher and parent report revealed the same *pattern of trajectories*, and predictors of those trajectories, for all three kinds of challenging behaviors. Consistency of results for parents and teachers further supports the idea that these identified patterns are accurate representations of participants’ behavior over time. It also provides evidence that behaviors (particularly hyperactivity and irritability) may often be relatively stable across home and school contexts, which involve different settings, situations, and social circumstances.

There was significant variability in hyperactivity, irritability, and social withdrawal trajectories based on various individual-level predictors. More specifically, participants with more autism features presented with more hyperactivity, irritability, and social withdrawal at age 9 and experienced greater improvement in these symptoms over time than participants with fewer ASD features. Similarly, participants with lower VIQs presented with more hyperactivity and irritability at age 9, though, in contrast to the effects of severity of autism behaviors, they experienced less improvement in hyperactivity, irritability, and social withdrawal across time compared to participants with higher VIQs. Interestingly, more slope effects (i.e., differences in rates of change based on VIQ or ASD features) were identified via teacher report than parent report; it may be that such individual differences matter more over time in the school setting than at home in which the environment remains more consistent. These findings from the present study are consistent with existing parent-report findings in the literature (Anderson et al., 2011; Shattuck et al., 2007; Woodman et al., 2015). Low VIQ among individuals with ASD and other developmental disabilities has been associated with increased risk for behavior problems (Beadle-Brown et al., 2006) and other negative developmental outcomes (Lord & Bailey, 2002; Seltzer et al., 2004).

The present study has important implications for clinical practice. Given the stable trajectory of social withdrawal and the impact of withdrawal on well-being and quality of life in ASD (Anderson et al., 2011; Kapp et al., 2011), promoting social approaches may be a particularly important intervention target during this developmental period. Additionally, despite the pattern of improvement in hyperactivity and irritability over time, interventions should continue to address these symptoms throughout development given the functional impairment caused by these symptoms (de Bildt et al., 2005; Kim et al., 2000). Furthermore, given that youth with more ASD features and lower VIQ demonstrated more challenging behaviors, providers should carefully assess for co-occurring challenging behaviors/conditions in patients with this diagnostic and cognitive profile and consider these co-occurring behaviors in intervention planning. The presence of challenging behaviors has been shown to exacerbate core deficits in ASD by hindering the development of social relationships (Matson & Wilkins, 2007; Matson et al., 2010), impeding placement in less restrictive environments (Shoham-Vardi et al., 1996), and limiting access to social interactions with peers (Jang et al., 2011; Matson et al., 2009). Thus, services targeting challenging behaviors may need to be prioritized to treat ASD-specific difficulties more effectively. Given the consistency in teacher- and parent-report patterns of challenging behaviors, these symptoms seem to emerge across settings. By supporting parents in managing challenging behaviors at home, it is possible that improvements in behaviors may have spill-over effects into school functioning and vice versa. Similarly, if a parent reports that their child is struggling with hyperactivity, irritability, or social withdrawal at home, it is likely that the child is also struggling with such behaviors at school. This highlights the need for multi-system interventions to improve functioning across settings.

In addition to the many strengths of this study including the use of a multi-informant approach and longitudinal design, there are limitations that highlight directions for future research. Children in our study were primarily referred for diagnosis under age 3; our findings may not be representative of children diagnosed with ASD at later ages. Additionally, characteristic of most longitudinal studies, attrition has affected the sample with increased participant dropout noted among participants from marginalized communities. Although the sample size is suitable for the present study, findings should be replicated with larger and more representative samples to bolster confidence in these conclusions. Additionally, the ABC has limitations inherent in informant report measures; future work may wish to include a clinical interview to provide more in-depth characterization of such challenging behaviors. Lastly, predictors of challenging behaviors that are outside the scope of the present study may be fruitful for future research to explore including, for example, intervention history and broader family-level factors.

## Conclusion

This longitudinal study aimed to examine trajectories of teacher- and parent-reported challenging behaviors, as well as predictors of these trajectories, from late childhood through early adulthood among individuals with ASD and non-spectrum delays. Over this timespan, results highlight a consistent pattern of improvement in hyperactivity and irritability, and minimal change in social withdrawal, across both parent and teacher reports in ASD. Higher cognitive ability and fewer autism features emerged as important individual differences related to fewer challenging behaviors. The multi-informant approach and longitudinal design provide novel insight into the manifestations of these challenging behaviors across different contexts and across time and can be used to inform clinical practice.

## Data Availability

Data produced in the present study are not available to others outside of the study team at this time.

## Acknowledgements

This work was supported by the National Institute of Child Health and Human Development [R01HD081199] (PI: C.L.) and the National Institute of Mental Health [R01MH081873] (PI: C.L.). The authors would like to thank the members of the Lord Lab for assistance with data collection and data entry. The authors would also like to thank the study participants and their families who made this research possible. Portions of this manuscript were presented at the 2022 Annual Meeting of the International Society for Autism Research (INSAR). C.L. acknowledges the receipt of royalties from the sale of the Autism Diagnostic Observation Schedule (ADOS). N.R. and H.S. have no potential conflicts to declare.

